# The iPAD study protocol: rationale, design and baseline characteristics of a randomised controlled trial of patient activation interventions for adults receiving chronic hemodialysis

**DOI:** 10.64898/2026.01.20.26344456

**Authors:** Edward Zimbudzi, Denise Fraginal-Hitchcock, Qiumian Wang, Lelise Gute, Asha Blessan, Shari Ziganay, Kevan R Polkinghorne

**Affiliations:** Department of Nephrology, Monash Health, Melbourne, Victoria, Australia; School of Nursing and Midwifery, Monash University, Melbourne, Victoria, Australia; Chronic and Complex Care Renal Services, Western Health, Melbourne, Victoria, Australia; School of Clinical Sciences, Monash University, Melbourne, Victoria, Australia; School of Public Health and Preventive Medicine, Monash University, Melbourne, Victoria, Australia

## Abstract

**Background:** Patient activation, defined as the knowledge, skills, and confidence to manage one’s health, is associated with better outcomes in chronic disease. However, evidence on interventions that improve activation in people with end-stage kidney disease on hemodialysis remains limited.

**Methods and analysis:** This single-centre, prospective, participant-blinded, randomised controlled trial conducted with adults undergoing chronic hemodialysis in an acute dialysis unit tests the hypothesis that adding tailored activation interventions to usual care improves patient activation and reduces complications in hemodialysis patients compared to usual care alone. A target sample size of 140 patients was recruited and randomised to iPAD interventions or usual care in a 1:1 ratio with an expected intervention period of at least 6 months. The primary outcome of iPAD was change in patient activation from baseline to 18 months.

**Ethics and dissemination:** This study has been approved by all institutional ethics review boards involved in the study. Participants could only be enrolled following informed written consent. Results will be published in peer-reviewed journals and presented at scientific and clinical conferences.

**Conclusion:** Recruitment and enrolment targets were successfully achieved, with the cohort broadly representative of the dialysis population, including strong participation from culturally and linguistically diverse and socioeconomically disadvantaged groups. The careful planning and successful execution of the study in resource-constrained environments highlight its feasibility and flexibility, establishing it as a scalable and cost-efficient model for broad implementation in dialysis care globally.

## Introduction

The growing prevalence of multiple chronic conditions, including kidney failure, poses a substantial challenge to healthcare systems both in Australia and globally [1–3]. In response, health systems are increasingly prioritising strategies aimed at improving care quality and outcomes. Central to these efforts is the recognition of the patient’s role in health management, with a strong emphasis on promoting active and effective self-management. This shift is associated with reduced unnecessary healthcare utilisation and improved health outcomes [4].

Patient activation, defined as an individual’s willingness and ability to take independent actions to manage their health and healthcare is a critical precursor to effective self-management [4]. It is widely recognised as a cornerstone of successful health system reform, as highly activated patients consistently yield better health outcomes [4, 5]. Activated patients understand their roles in the care process and are equipped with the necessary knowledge, skills and confidence to manage their health. On the other hand, individuals with low activation are likely to have less disease knowledge [6], increased likelihood to visit the emergency department, unhealthy habits or poor lifestyle such as smoking [7], low health related quality of life [8] and less favourable care experiences compared to those with higher activation levels [9].

Several studies have shown that patient activation is generally low among individuals with moderate to severe chronic kidney diseases (CKD) (stages 3 and 4) [10, 11] and significantly lower in those with ESKD [11, 12]. While numerous studies have examined the impact of patient activation interventions in individuals with CKD, primarily to support blood pressure control and slow CKD progression, such interventions have not been evaluated in populations with advanced CKD [13–17]. The limited evidence base regarding the benefits of patient activation among individuals with ESKD, particularly those receiving chronic hemodialysis, underscores the need for prospective research examining its relationship with clinical outcomes, including mortality [18]. Outcomes related to patient activation in this population remain underexplored. To advance patient-centred, value-based care and move away from paternalistic models of care [19], it is essential to develop and evaluate and utilise activation interventions tailored to individuals with ESKD.

Interventions aimed at enhancing patient activation among individuals receiving chronic hemodialysis have the potential to yield significant clinical benefits and improvements in patient-reported outcomes. An ideal activation-focused intervention would support optimal management of inter-dialytic weight gain (IDWG), blood pressure, medication adherence, and dietary behaviours. Most importantly, such an intervention may lead to meaningful improvement in health-related quality of life for people undergoing chronic hemodialysis. To optimise outcomes, patient activation interventions should adopt a holistic and tailored approach. This includes engaging family support systems, addressing social determinants of health, and they should be guided by patient-identified goals and priorities.

We report baseline data from a randomised controlled trial evaluating the impact of a nurse-led, tailored, person-centred intervention on patient activation and clinical outcomes in individuals undergoing hemodialysis.

## Methods

### Study design and setting

The **I**nterventions to support **P**atient **A**ctivation for adults on hemo**D**ialysis (iPAD) study (ACTRN12619000504112) is a single-centre, prospective, participant-blinded, randomised controlled trial conducted among adults receiving chronic hemodialysis at Monash Medical Centre, a tertiary care hospital in Victoria, Australia.

### Recruitment and eligibility

Participant recruitment took place between March 2018 and October 2023 and follow is in progress. Participants were eligible for inclusion if they were aged 18 years or older, had commenced hemodialysis within the preceding three months, and were fluent in English. Exclusion criteria included cognitive impairment, a diagnosis of acute kidney injury, plans to permanently transfer to a hemodialysis unit outside the public health system, or having been on hemodialysis for more than three months (Figure 1). All eligible participants received study information and provided written informed consent prior to enrolment. The study was reviewed and approved by the Monash Health Human Research Ethics Committee (Approval number: RES-17-0000-577A) and was conducted in accordance with the National Statement on Ethical Conduct in Human Research [20].

**Figure 1.**
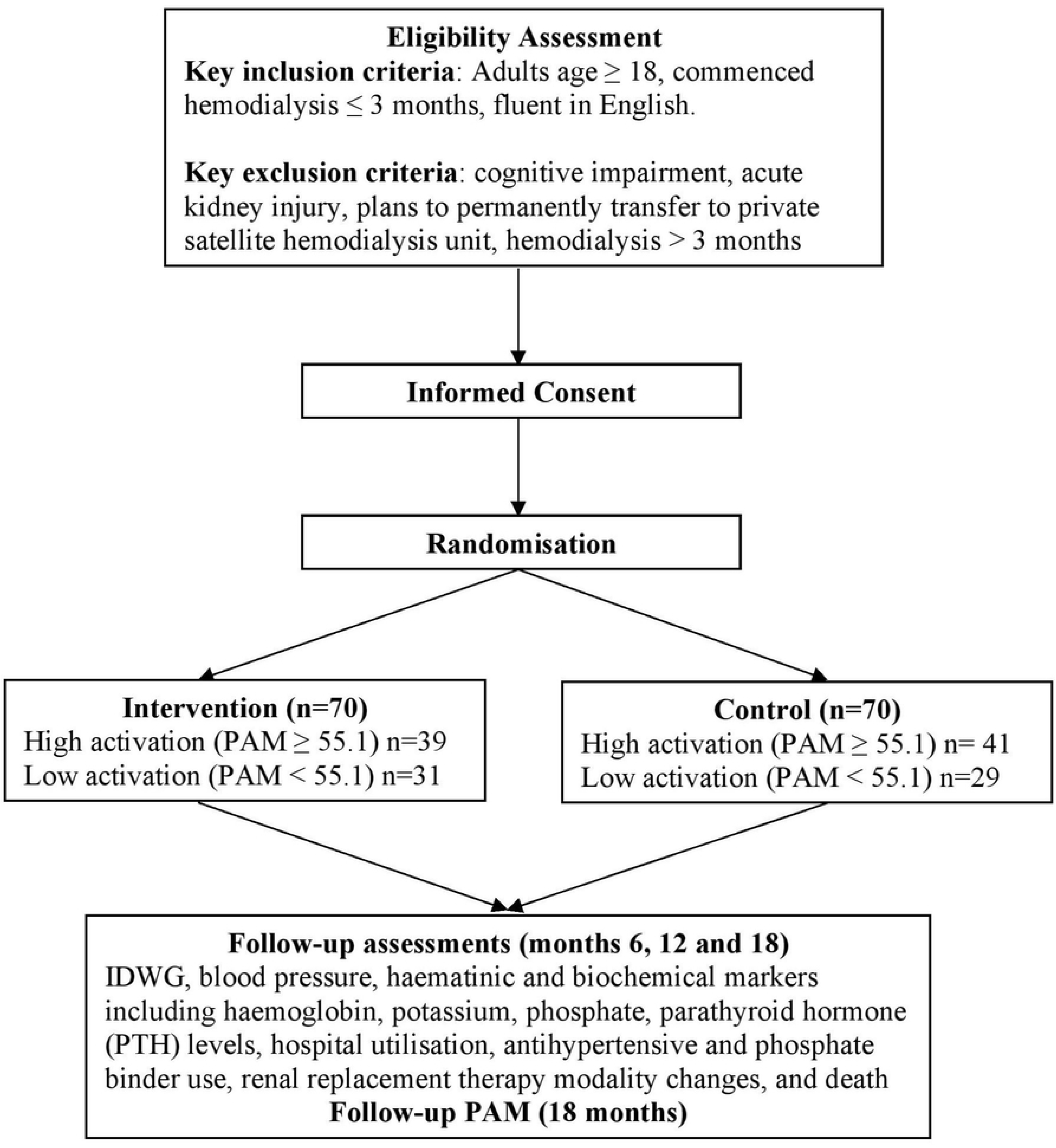
IPAD study flowchart.

**Figure 2.**
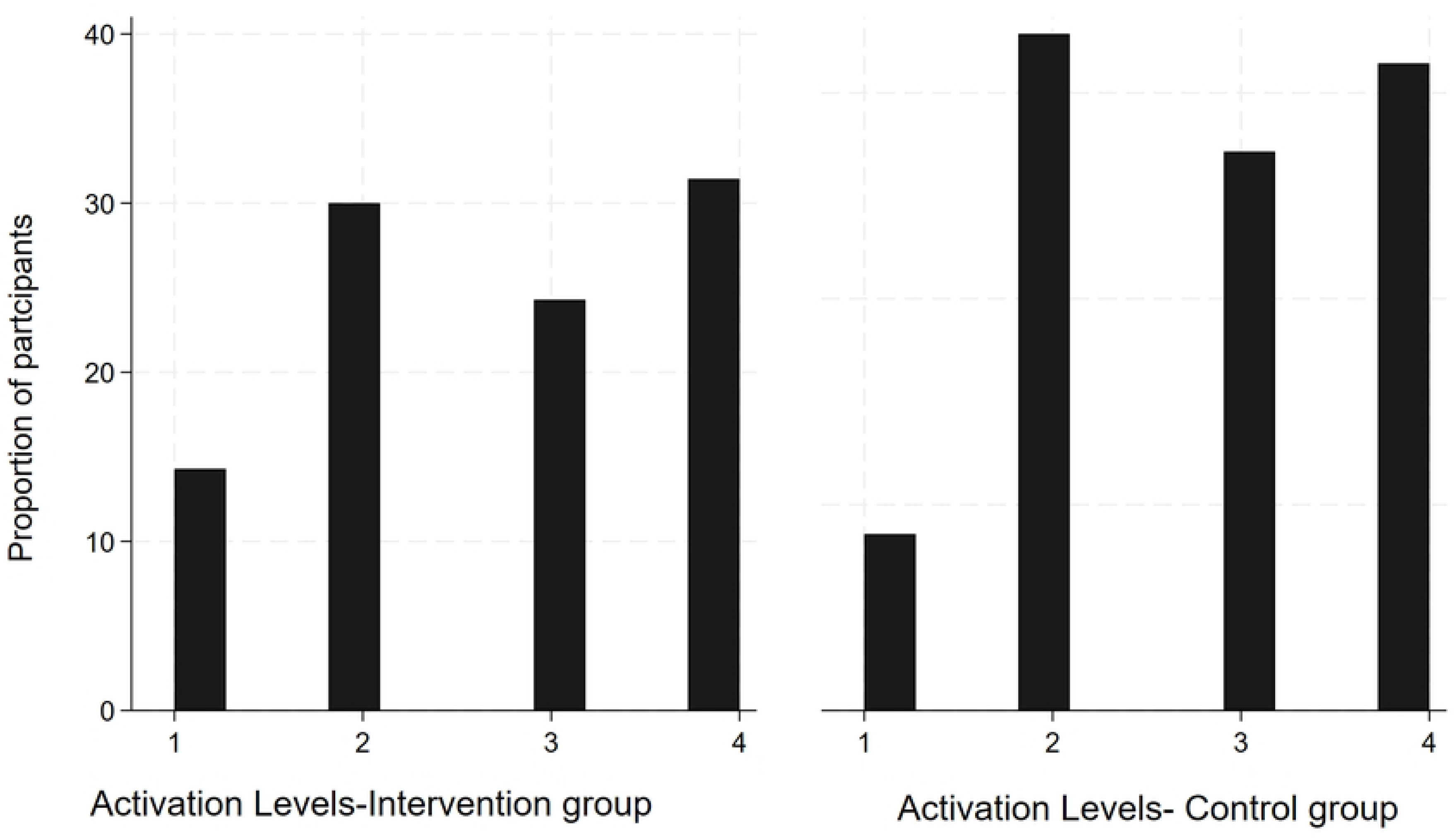
Distribution of patients across levels of activation.

### Randomisation

Random allocation was accomplished using a concealed, random number list prepared by an independent clinician with randomly permuted block sizes, stratified by baseline Patient Activation Measure (PAM) score (≥55.1 indicating high activation; <55.1 indicating low activation), using Stata statistical software.

#### Intervention Group (IPAD Program)

The iPAD program is a six-month, tailored intervention designed to enhance patient activation and self-management among individuals with CKD (details of the intervention are summarised in Table 1). The intervention was stratified according to baseline activation levels, classified as either low or high, to ensure appropriate intensity and support. All participants received a one-hour face to face, individualised self-management training session in Week 1, delivered by nephrology nurses trained in motivational interviewing and self-management support. The session introduced the concept and benefits of self-management, provided education on dialysis, medication management, and lifestyle adjustments, and facilitated the development of a personalised action plan. Participants were also provided with a logbook to monitor blood pressure, fluid intake, daily weight, medications, and diet. Those in the high activation group additionally received standardised educational materials from Kidney Health Australia addressing common concerns such as hyperkalemia, fluid management, and hyperphosphatemia.

**Table 1:** Schedule of the iPAD intervention.

| Time | Intervention design |  |
| --- | --- | --- |
|  | Interventions for Low Activated Group | Interventions for High Activated Group |
| Week 1 | *Individual self-management training-1 hour of one-on-one session* delivered by trained nephrology nurses with motivational interviewing and self-management support training.<br>Content:<br>• introduction to the self-management concept<br>• addressing the importance and benefits of self-management<br>• providing education around dialysis, medication management and lifestyle adjustment required<br>• forming an action plan based on participant-identified health related concern, introducing a logbook to record blood pressure, fluid intake, daily weight, medications and diet. | *Individual self-management training-1 hour of one-on-one session* delivered by trained nephrology nurses with motivational interviewing and self-management support training.<br>Content:<br>• forming an action plan based on participant-identified health related concern, introducing a logbook to record blood pressure, fluid intake, daily weight, medications and diet<br>• distribution of standard pamphlets and resources from Kidney Health Australia providing information on common concerns like hyperkalaemia, fluid management, hyperphosphatasemia |
| Week 2-5 | *Feedback and tailored support-weekly telephone follow-ups* for one month after the initial training.<br>Content: | *Feedback and tailored support-fortnightly telephone follow-ups* for one month.<br>Content: |
|  | • Phone calls focused on participant-highlighted concern and tailored support provided on how to manage this concern.<br>• Formulation of participant-led care plans<br>• Encouragement for questions and active participation.<br>• Additional phone calls from the study nurses prior to clinic appointments to formulate questions for the clinician in order to maximise the value of the appointment and promote participation in decision-making. | • Phone calls focused on participant-highlighted concern and tailored support provided on how to manage this concern.<br>• Formulation of participant-led care plans<br>• No additional phone calls prior to clinic appointments; however, follow up with regards to outcome of clinic appointments was sought from the participants in this subgroup |
| Week 6-9 | *Feedback and tailored support- **fortnightly telephone follow-ups** for one month.*<br>Content:<br>• Focus on participant-highlighted concern and tailored support provided on how to manage this concern.<br>• Encouraging for questions and active participation.<br>• Updating of care plans | *Feedback and tailored support-**monthly telephone follow-ups** for one month.*<br>Content:<br>• Focus on participant-highlighted concern and tailored support provided on how to manage this concern.<br>• Updating of care plans |
| Week 10- 26 | *Feedback and tailored support- **monthly telephone follow-ups** up to the 6<sup>th</sup> month for participants.*<br>Content:<br>• Focus on participant-highlighted concern and tailored support provided on how to manage this concern.<br>• Encouraging for questions and active participation.<br>• Updating of care plans | No further interventions |

**Low Activation Group:** Participants received intensive follow-up support:

- Weeks 2–5: Weekly telephone calls focused on participant-identified concerns, development of participant-led care plans, and preparation for clinic visits to enhance shared decision-making.
- Weeks 6–9: Fortnightly telephone calls emphasised care plan updates and active engagement.
- Weeks 10–26: Monthly telephone calls provided ongoing support for the remainder of the intervention period.

**High Activation Group:** Participants received a lighter-touch follow-up schedule:

- Weeks 2–5: Fortnightly telephone calls addressed participant concerns and supported care plan development.
- Weeks 6–9: Monthly calls focused on updating care plans.
- No further intervention was provided beyond Week 9, although participants were asked to report outcomes from clinic visits.

#### Control Group

Participants in the control group received routine care consistent with standard practice for patients undergoing hemodialysis. This included education and support from healthcare providers such as nephrologists, dialysis educators, nephrology nurses, renal dietitians, and social workers. Standard care encompassed information on the hemodialysis process, symptom management, medication adherence, dietary guidance, fluid restriction, and psychosocial coping strategies. Educational materials were delivered through standardised pamphlets and electronic resources

#### Outcomes

##### Primary Outcome

The primary outcome was change in patient activation from baseline to 18 months. Patient activation was assessed using the 13-item short form of the Patient Activation Measure (PAM-13), which categorises individuals into four levels according to their degree of activation. The PAM-13 demonstrates reliability and validity comparable to the original 22-item version across diverse ages, genders, and health conditions, with a Cronbach’s alpha of 0.91 and Rasch person reliability statistics of 0.81 (real) and 0.85 (model-based) [21, 22]. Its psychometric properties have also been validated in various regions and among patients with a range of health conditions [23, 24]. Each item offers five response options scored from 0 to 4: (0) not applicable, (1) disagree strongly, (2) disagree, (3) agree, and (4) agree strongly. The total raw score ranges from 13 to 52 and is converted to a standardised activation scale ranging from 0 to 100, following PAM scoring guidelines. Participants were classified into one of four activation levels: Level 1 (least activated) with scores ≤47; Level 2 with scores between 47.1 and 55.1; Level 3 with scores between 55.2 and 67; and Level 4 (most activated) with scores ≥67.1. PAM assessments were conducted at baseline and at 18 months follow-up (**Table 2**). The choice to measure PAM at just two time points was deliberate to reduce recall bias. This type of bias occurs when participants remember their earlier responses or test questions, which can impact their answers in later assessments. By limiting the frequency of testing, the study aims to ensure that participants’ responses reflect their actual condition rather than being influenced by memory, thereby improving the reliability and validity of the findings.

**Table 2:** Schedule of Baseline and Follow-Up Outcome Measures.

| Outcomes | Time point (months) |  |  |  |
| --- | --- | --- | --- | --- |
|  | 0 | 6 | 12 | 18 |
| PAM | ✓ | - | - | ✓ |
| Interdialytic weight gain | ✓ | ✓ | ✓ | ✓ |
| Blood pressure | ✓ | ✓ | ✓ | ✓ |
| Haemoglobin | ✓ | ✓ | ✓ | ✓ |
| Potassium | ✓ | ✓ | ✓ | ✓ |
| Phosphate | ✓ | ✓ | ✓ | ✓ |
| Parathyroid hormone | ✓ | ✓ | ✓ | ✓ |
| Antihypertensive use | ✓ | ✓ | ✓ | ✓ |
| Phosphate binder use | ✓ | ✓ | ✓ | ✓ |
| Emergency Department presentation | ✓ | ✓ | ✓ | ✓ |
| Hospital admission | ✓ | ✓ | ✓ | ✓ |

**Table 3.** Baseline characteristics of participants in the IPAD study.

| Characteristic | All | Intervention | Control |
| --- | --- | --- | --- |
| | Mean ± SD, n (%) | | |
| Age, years (mean ± SD) | 58.4 ± 14.8 | 59 ± 15 | 58 ± 15 |
| Male | 101 (72.1) | 52 (74) | 49 (70) |
| Country of birth-Australia and NZ | 71 (50.7) | 31 (44.3) | 37 (52.9) |
| Married | 80 (57.1) | 36 (51.4) | 44 (62.9) |
| Education-up to year 12 | 82 (58.6) | 42 (60.9) | 40 (57.1) |
| Employed | 42 (30.0) | 18 (25.7) | 25 (34.3) |
| Primary carer – Self | 92 (65.7) | 45 (64.3) | 47 (67.1) |
| Socioeconomic status |  |  |  |
| Upper | 51 (36.4) | 26 (37.1) | 25 (35.7) |
| Upper middle | 15 (10.7) | 6 (8.6) | 9 (12.9) |
| Lower middle | 32 (22.9) | 18 (25.7) | 14 (20) |
| Upper lower | 11 (7.9) | 4 (5.7) | 7 (10) |
| Lower | 31 (22.1) | 16 (22.9) | 15 (21.4) |
| **Primary disease** |  |  |  |
| Diabetes | 58 (41.4) | 31 (44.3) | 27 (38.6) |
| Glomerulonephritis | 28 (20) | 11 (15.7) | 17 (24.3) |
| Hypertension | 11 (7.9) | 6 (8.6) | 5 (7.1) |
| Diabetes/Hypertension | 4 (2.9) | 2 (2.9) | 2 (2.9) |
| Glomerulonephritis | 7 (5.0) | 5 (7.1) | 2 (2.9) |
| Polycystic Kidney Disease | 7 (5.0) | 2 (2.9) | 5 (7.1) |
| Reflux | 3 (2.1) | 2 (2.9) | 1 (1.4) |
| Other | 22 (15.7) | 11 (15.7) | 11 (15.7) |
| **Clinical characteristics** |  |  |  |
| Serum potassium, mmol/L | 4.6 ± 0.6 | 4.6 ± 0.5 | 4.6 ± 0.6 |
| Phosphate, mmol/L | 1.7 ± 0.6 | 1.7 ± 0.6 | 1.8 ± 0.6) |
| Parathyroid hormone, pg/mL | 42.5 ± 33.5 | 41.1 ± 37.4 | 44.7 (29.0) |
| Hemoglobin, g/L | 98.3 ± 15.1 | 96 ± 14.7 | 100.5 ± 15.2 |
| Average interdialytic weight gain, kg | 1.7 ± 0.9 | 1.6 ± 1.0 | 1.8 ± 0.9 |
| Systolic blood pressure, mmHg | 151.0 ± 22.3 | 152.7 ± 24.2 | 149.2 ± 20.2 |
| Diastolic blood pressure, mmHg | 76.8 ± 15.9 | 78 ± 16.3 | 75.6 ± 15.6 |
| Patients taking phosphate binders | 83 (59.3) | 40 (57) | 43 (61) |
| Patients taking antihypertensives | 126 (90.0) | 61 (87) | 65 (93) |
| Co-morbidity index |  |  |  |
| Mild comorbidity | 18 (12.9) | 8 (11.5) | 10 (14.3) |
| Moderate comorbidity | 44 (31.4) | 26 (37.1) | 18 (25.7) |
| Severe comorbidity | 78 (55.7) | 36 (51.4) | 42 (60) |
| PAM score | 61.9 ± 15.0 | 61.3 ± 14.3 | 62.5 ± 15.7 |

SD-standard deviation; NZ-New Zealand; PAM-Patient Activation Measure; FSGS- Focal Segmental Glomerulosclerosis

##### Secondary Outcomes

Secondary outcome measures included IDWG, blood pressure, haematinic and biochemical markers including haemoglobin, potassium, phosphate, parathyroid hormone (PTH) levels, hospital utilisation, antihypertensive and phosphate binder use, renal replacement therapy modality changes, and death. These measures were evaluated at baseline and then every six months for a total of 18 months (**Table 2**).

### Sample size estimation

Previous studies suggest the minimal clinically important difference for the PAM is 4 to 5.4 [25, 26]. Assuming a mean of 60, as previously reported in this population [27, 28], and standard deviation of 14 in the control group, we estimated that 63 participants in each group would be needed detect an 8-point difference in mean PAM scores between the intervention and control groups. Allowing for a 10% attrition rate, a total sample size of 140 participants (70 in each group) was determined.

### Statistical Analysis

All analyses will be conducted on an intention-to-treat basis, with a per-protocol analysis performed as a sensitivity analysis. Baseline characteristics will be summarised using means and standard deviations for normally distributed continuous variables, medians and interquartile ranges for skewed data, and counts and percentages for categorical variables; no formal hypothesis testing will be conducted for baseline balance, although standardised differences will be reported. The primary outcome, change in PAM score from baseline to the primary follow-up, will be analysed using analysis of covariance (ANCOVA) with adjustment for baseline PAM scores. Results will be presented as mean difference with 95% confidence intervals. Missing data will be addressed using multiple imputation by chained equations under a missing-at-random assumption, with complete-case and other sensitivity analyses to assess robustness. Secondary continuous outcomes will be analysed similarly to the primary outcome, binary outcomes will be assessed using logistic regression, and time-to-event outcomes will be evaluated using Kaplan–Meier methods and Cox proportional hazards models. Exploratory subgroup analyses, using treatment-by-subgroup interaction terms, will be performed for age, sex, baseline PAM category. Adverse events will be summarised by treatment group and compared using Fisher’s exact or chi-square tests as appropriate. Statistical significance will be set at p < 0.05 for the primary outcome, with secondary outcomes interpreted as exploratory. Analyses will be performed using R and Stata statistical software.

### Progress

A total of 147 participants were screened for eligibility, with 140 consenting to participate in the trial. Recruitment took place predominantly during the height of COVID-19 restrictions in Melbourne, which included multiple periods of stringent lockdown. Despite these challenging circumstances, recruitment efforts continued uninterrupted; however, the pace of enrolment was notably slowed. Several factors contributed to this reduction, including limited staffing levels as clinical research personnel were reallocated to essential healthcare duties, and logistical challenges associated with maintaining research activities during the pandemic. Importantly, the intervention was consistently delivered through face-to-face sessions throughout the study period. The research team deliberately chose not to transition to a virtual delivery format to preserve the integrity and uniformity of the intervention experience for all participants. This approach aimed to minimise variability in intervention exposure and ensure comparability across study arms.

### Baseline Characteristics

The study cohort had a mean age of 58.4 years (±14.8), with males comprising 72.1% (n = 101) of participants. Just over half of the participants (50.7%, n = 71) were born in Australia or New Zealand. Most participants were married (57.1%, n = 80), and 58.6% (n = 82) had education up to Year 12. Employment was reported in 30.0% (n = 42) of participants, and 65.7% (n = 92) identified as their own primary carer. Socioeconomic status, classified according to the Socio-Economic Indexes for Areas, was distributed with 36.4% (n = 51) in the upper category, 10.7% (n = 15) in upper middle, 22.9% (n = 32) in lower middle, 7.9% (n = 11) in upper lower, and 22.1% (n = 31) in the lower category.

Diabetes was the most common primary cause of kidney failure, affecting 41.4% (n = 58) of participants, followed by IgA nephropathy (15.0%, n = 21) and hypertension (7.9%, n = 11). Clinical parameters indicated a mean serum potassium level of 4.6 mmol/L (±0.6), phosphate of 1.7 mmol/L (±0.6), parathyroid hormone concentration of 42.5 pmol/L (±33.5), and hemoglobin level of 98.3 g/L (±15.1). The average interdialytic weight gain was 1.7 kg (±0.9). Blood pressure readings showed a mean systolic pressure of 151.0 mmHg (±22.3) and diastolic pressure of 76.8 mmHg (±15.9). The majority of participants were prescribed antihypertensives (90.0%, n = 126) and phosphate binders (59.3%, n = 83).

Comorbidity burden was substantial, with 12.9% (n = 18) classified as having mild, 31.4% (n = 44) moderate, and 55.7% (n = 78) severe comorbidity based on the comorbidity index. The mean PAM score at baseline was 61.9 (±15.0), indicating moderate levels of patient engagement in self-management.

## Discussion

Recruitment to this trial presented both challenges and opportunities. The narrow eligibility window, limiting participation to individuals within three months of commencing haemodialysis meant that the recruitment pool was inherently small. Although this approach necessitated more rigorous screening and coordination, it ensured the recruitment of a clinically relevant and homogeneous patient cohort, thereby enhancing the internal validity of the study. Moreover, restricting enrolment to patients within the first three months of dialysis initiation allowed for a more accurate assessment of the intervention’s true effect by minimising contamination from concurrent interventions and educational exposures over time. Future studies in similar contexts may build on this approach by adopting multi-site recruitment or implementing rolling enrolment strategies to sustain momentum while preserving sample relevance.

The COVID-19 pandemic inevitably influenced recruitment dynamics, but it also prompted innovation and resilience in trial conduct. Lockdowns, workforce redeployments, and heightened infection control protocols created logistical hurdles at our recruitment site [29], mirroring challenges reported in trials worldwide [30]. Despite these constraints, recruitment continued without full suspension, reflecting the commitment of both participants and clinical teams. This is consistent with emerging evidence indicating that trials embedded within active clinical networks and employing stakeholder-driven approaches are better positioned to withstand external disruptions [31]. The experience reinforces the value of building strong site-level collaborations and flexible staffing models as part of trial preparedness.

Importantly, the decision to retain in-person intervention delivery, rather than transitioning to a remote format, helped maintain fidelity and participant engagement. While virtual delivery can enhance reach, particularly during crises, literature suggests that face-to-face interactions in complex behavioural or educational interventions can strengthen rapport and adherence [32]. The trial’s ability to continue under pandemic conditions without compromising intervention integrity demonstrates that with careful planning and team coordination, high-quality RCTs can be executed even in highly challenging circumstances. This adaptability provides a framework for future research seeking to balance methodological rigour with operational flexibility.

Despite recruitment delays caused by the COVID-19 pandemic, the iPAD study represents the first global trial to investigate the effect of tailored interventions on patient activation in dialysis patients. Designed prior to the pandemic, this trial is poised to demonstrate that delivering such interventions is feasible even in resource-constrained environments. This adaptability positions the iPAD intervention as a scalable model suitable for adoption in dialysis units worldwide. Moreover, as an investigator-initiated study conducted without external funding, iPAD underscores the feasibility and applicability of implementing effective interventions with minimal resources.

### Patient and public involvement

Patients and public will not be involved in the design, or conduct, or reporting, or dissemination plans of our study.

## Data Availability

All relevant data are within the manuscript and its Supporting Information files

